# Comparative Analysis of Cytokine Profiles in Cerebrospinal Fluid and Blood Serum in Patients with Acute and Subacute Spinal Cord Injury

**DOI:** 10.1101/2023.08.29.23294725

**Authors:** Davran Sabirov, Sergei Ogurcov, Ilya Shulman, Ilyas Kabdesh, Ekaterina Garanina, Albert Sufianov, Albert Rizvanov, Yana Mukhamedshina

## Abstract

**Background:** Cytokines are actively involved in the regulation of the inflammatory and immune responses and play crucial importance for the outcome of spinal cord injury (SCI). Considering the importance of examining more objective and representative indicators of the patient’s condition is still required to reveal the fundamental patterns of abovementioned posttraumatic processes, including identification of changes in the expression of cytokines.

**Methods:** We performed a dynamic (3, 7 and 14 dpi) extended multiplex analysis of cytokine profiles in both CSF and blood serum of SCI patients with baseline American Spinal Injury Association Impairment Scale grades of A.

**Results:** The data obtained showed a large elevation of IL-6 (>58 fold) in CSF and IFN-γ (>14 fold) in blood serum at 3 dpi with a downward trend as the post-traumatic period increases. The level of cytokine CCL26 was significantly elevated both CSF and blood serum at 3 days post-SCI while other cytokines did not show the same trend in the different biosamples.

**Conclusions:** The dynamic changes in cytokine levels observed in our study can explore the relationships with SCI region and injury severity, paving the way for a better understanding of the pathophysiology of SCI and potentially more targeted and personalized therapeutic interventions.

## 1. Introduction

Spinal cord injury (SCI) is a global health problem affecting tens of thousands of people every year, with profound consequences for those affected and significant societal costs. Neurological disorders resulting from SCI often lead to severe disability and dramatically change the lives of patients and their families (Ahuja, Wilson, et al., 2017). While SCI used to predominantly affect younger individuals, its incidence is now equally high among the elderly (Singh et al., 2014).

Despite significant advances in medical, surgical, and rehabilitation care for people with SCI, measures and interventions to ensure neurological recovery remain limited. This limited recovery is primarily due to the central nervous system’s (CNS) inherently low regenerative capacity, the complexity of the injury, and poorly understood pathophysiological events after SCI (Ahuja, Nori, et al., 2017).

Evaluating new treatments for SCI presents a significant challenge. Demonstrating neurological recovery during the acute phase is particularly challenging due to the potential for spontaneous recovery (Fehlings et al., 2017). This problem is exacerbated by the fact that in the clinical evaluation of new treatments, we rely on a functional neurological examination only (S. C. Kirshblum et al., 2011). Although tools such as the International Standards for Neurological Classification of Spinal Cord Injury (ISNCSCI) provide a standardized assessment of neurological function, it has limitations when used in clinical trials (S. Kirshblum et al., 2021). This highlights the importance of examining other, more objective and representative indicators of the patient’s condition, such as biomarkers, for example.

In light of the need to search for more reliable and stable biomarkers in SCI, it seems mor e representative to study the cerebrospinal fluid (CSF), which is in close contact with the CNS and can provide a more accurate picture of the activity of various molecules in the area of injury. However, it should be noted that blood sampling is more acceptable and safer, and blood serum analysis can also provide information on some key biomarkers, although it requires additional research to clarify its value as a possible diagnostic and prognostic tool in the context of SCI.

Analysis of changes in cytokine levels at different periods after injury and their correlation with the American Spinal Injury Association (ASIA) Impairment Scale score can provide valuable insights not only into the pathophysiology of SCI, but also potentially guide therapeutic interventions (Biglari et al., 2015; Y. Chen et al., 2020). By including a more samples of SCI patients and using multiplex analysis, our aim was to determine how cytokine change correlates with time after severe injury, which may help develop new strategies for early diagnosis and personalized treatment of SCI.

## 2. Materials and Methods

### 2.1. Study population

All patients (n=40) were recruited at the Neurosurgical Department No. 2 of the Republican Clinical Hospital (Kazan, Russia). Written informed consent was obtained from each subject before CSF and blood serum samples were obtained. This study was approved by the Kazan Federal University Local Ethical Committee (Protocol No. 3, 23 March 2017).

The following inclusion criteria were used for patients with acute and subacute SCI in this prospective trial: (1) over 18 years of age; (2) SCI between C3 and L3 inclusive; (3) the ability to provide a valid, reliable neurological examination; (4) classified with an ASIA grade A (motor and sensory complete paralysis) SCI upon presentation; (5) no concomitant traumatic brain injury, severe chest injuries, and intra-abdominal injuries. The severity of neurological impairment with baseline ASIA grade assessments were conducted by a research study neurologist experienced in these techniques and in calculating ASIA. Neurological examinations were additionally conducted at 1- and 2-weeks post-injury to determine ASIA conversion and possible motor and sensory score improvement. Among the SCI patients, injuries were located in the cervical (C) region for 18 patients, thoracic (Th) for 14 patients, and lumbar (L) for 8 patients. Of the 40 patients, 35 (87%) were male, and the mean age was 42.2±15.1 years.

The blood serum samples for uninjured control group were collected from 16 healthy able-bodied individuals, from whom we obtained consent to acquire venous blood. Uninjured subjects were recruited from the Kazan Federal University student and academic population, and hospital staff. The CSF samples were collected from conditionally healthy patients who sought medical attention due to lumbar disc herniations or stenosis, from whom we obtained written informed consent. The following inclusion criteria were used for uninjured subjects: (1) over 18 years of age; (2) without a history of SCI or brain injury; (3) a lack of the main signs of acute or chronic inflammation and autoimmune processes; and (4) a normal complete blood count.

### 2.2. Samples collection and storage

The CSF and venous blood samples were obtained from SCI patients at 3, 7 and 14 days post-injury (dpi). The results of blood serum multiplex analysis at 14 dpi were presented by us earlier (Ogurcov et al., 2021). The CSF samples from uninjured control group were collected the day before lumbar spine surgery. Venous blood was collected via standard venipuncture in 6 mL vacuum test tube (Apexlab, Moscow, Russia). After 30 min of coagulation, the blood was centrifuged at 3000 RPM, divided into aliquots (300 µL), and stored at −80 ◦C until analysis.

The CSF samples were collected following strict aseptic technique, a lumbar puncture with an atraumatic needle (Medispine, 20G, India) was performed at L3-4 and a 3 mL sample of CSF was collected. The CSF samples were divided into 500 µl aliquots, centrifuged at 1000 RPM for 10 min, and the supernatant then immediately frozen and stored at −80°C. All obtained samples from injured patients and uninjured control group were subjected to the same manipulations and similar storage times.

It is important to note that some patients with SCI may develop cerebrospinal fluid dynamics disorders, which can make it difficult to collect cerebrospinal fluid at various stages after injury. Because of this, we were unable to obtain data for all 40 patients at investigated time points after injury. This means that the number of samples used in the analysis may vary depending on the time of collection.

### 2.3. Multiplex Analysis

In our study, we analyzed changes in the cytokine profile of CSF and blood serum of SCI patients at various dpi using multiplex analysis using the technology by xMAP Luminex. Bio-Plex Pro™ #171AK99MR2 (Bio-Rad, CA, USA) was used, allowing simultaneous multiplex analysis of 40 human cytokines (CCL21, CXCL13, CCL27, CXCL5, CCL11, CCL24, CCL26, CX3CL1, CXCL6, GMCSF, CXCL1, CXCL2, CCL1, IFN-γ, IL1b, IL2, IL4, IL-6, CXCL8/IL-8, IL10, IL16, CXCL10, CXCL11, MCP-1, MCP-2/CCL8, MCP-3, MCP-4, CCL22, MIF, MIG/CXCL9, MIP-1a/CCL3, MIP-1b/CCL4, MIP-3a/CCL20, MIP-3b/CCL19, MPIF-1/CCL23, CXCL16, SDF-1/CXCL12, CCL17, CCL25 и TNFα) in 50 µL of the test sample. All obtained blood serum samples were analyzed together in the same assay.

### 2.4. Statistical Analysis

We used R 3.6.3 (R Foundation for Statistical Computing, Vienna, Austria) for data analysis. Descriptive statistics for quantitative variables are presented as mean (standard deviation) and median (interquartile range). Prior to test selection, data were assessed for normality with the D’Agostino Pearson test. The Kruskal–Wallis test was used to test for an overall difference in median levels of cytokines between groups while using the Dunn’s correction for multiple comparisons. The Benjamini–Hochberg procedure was applied for multiplicity correction.

## 3. Results

### 3.1. Dynamics of cerebrospinal fluid cytokine profile after spinal cord injury

We analyzed 40 cytokines in the CSF of 35 patients at various days post-SCI, revealing significant changes in cytokine concentrations at 3, 7, and 14 dpi (Figure 1). In particular, we found a significant increase in the levels for CCL22 (2.7-fold, P.adj < 0.009), CCL26 (6.6-fold, P.adj < 0.017), IL-8 (6.7-fold, P.adj < 0.013), CCL23 (8.2-fold, P.adj < 0.04) and IL-6 (58.8-fold, P.adj < 0.001) at 3 dpi compared to uninjured control samples (Figure 2, Supplementary Table S1). At 7 and 14 dpi we continued to observe elevated levels of the abovementioned cytokines, although their concentrations were somewhat lower than at 3dpi.

**Figure 1.**
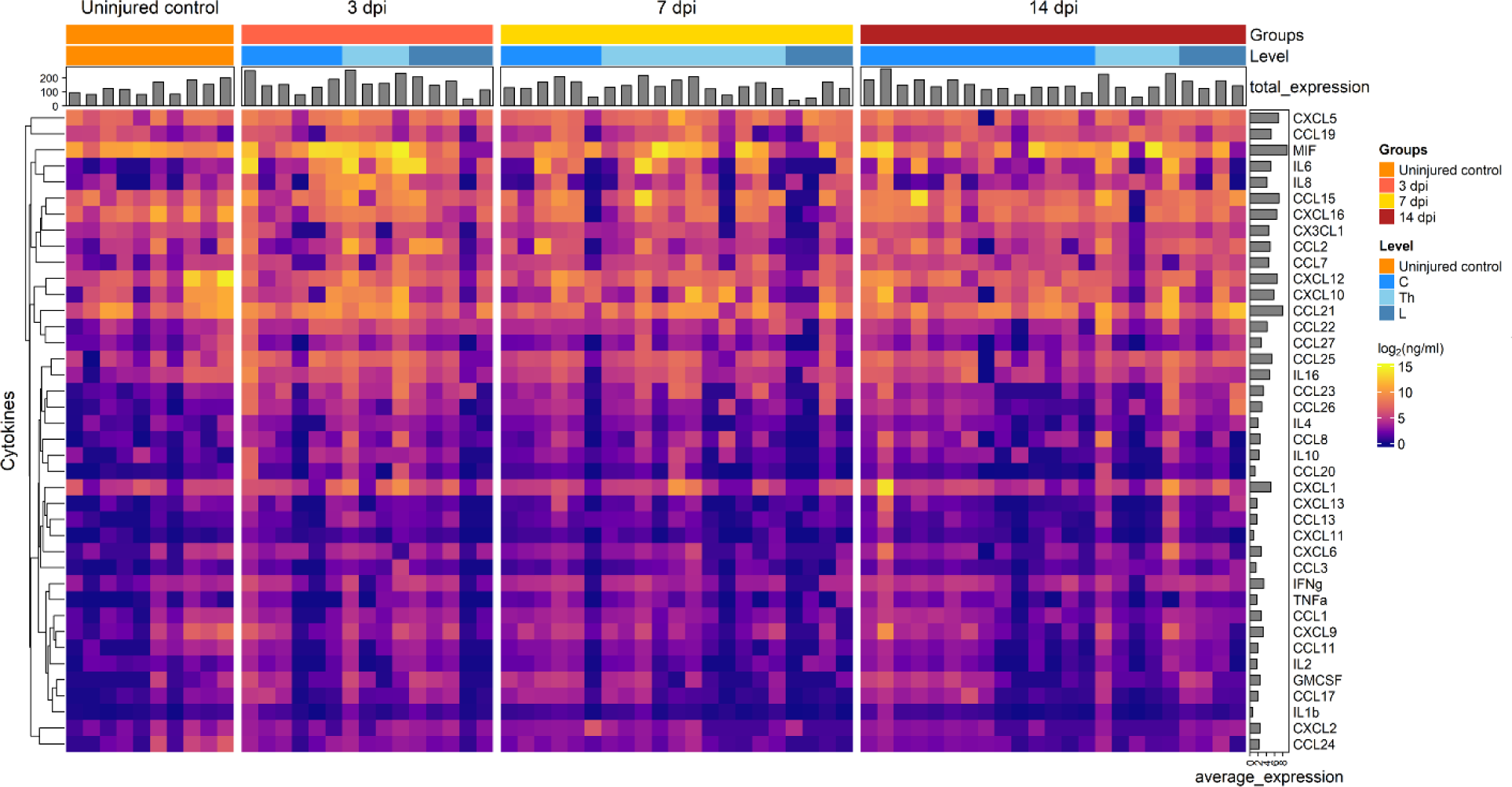
Graphical representation showing log2 cytokine concentrations (color keys), generated with the multiplex analysis of the CSF collected at 3dpi (n=15), 7dpi (n=21), 14dpi (n=23) or from uninjured control (n=10). A dendrogram resulting from hierarchical clustering of cytokines is shown on the left.

**Figure 2.**
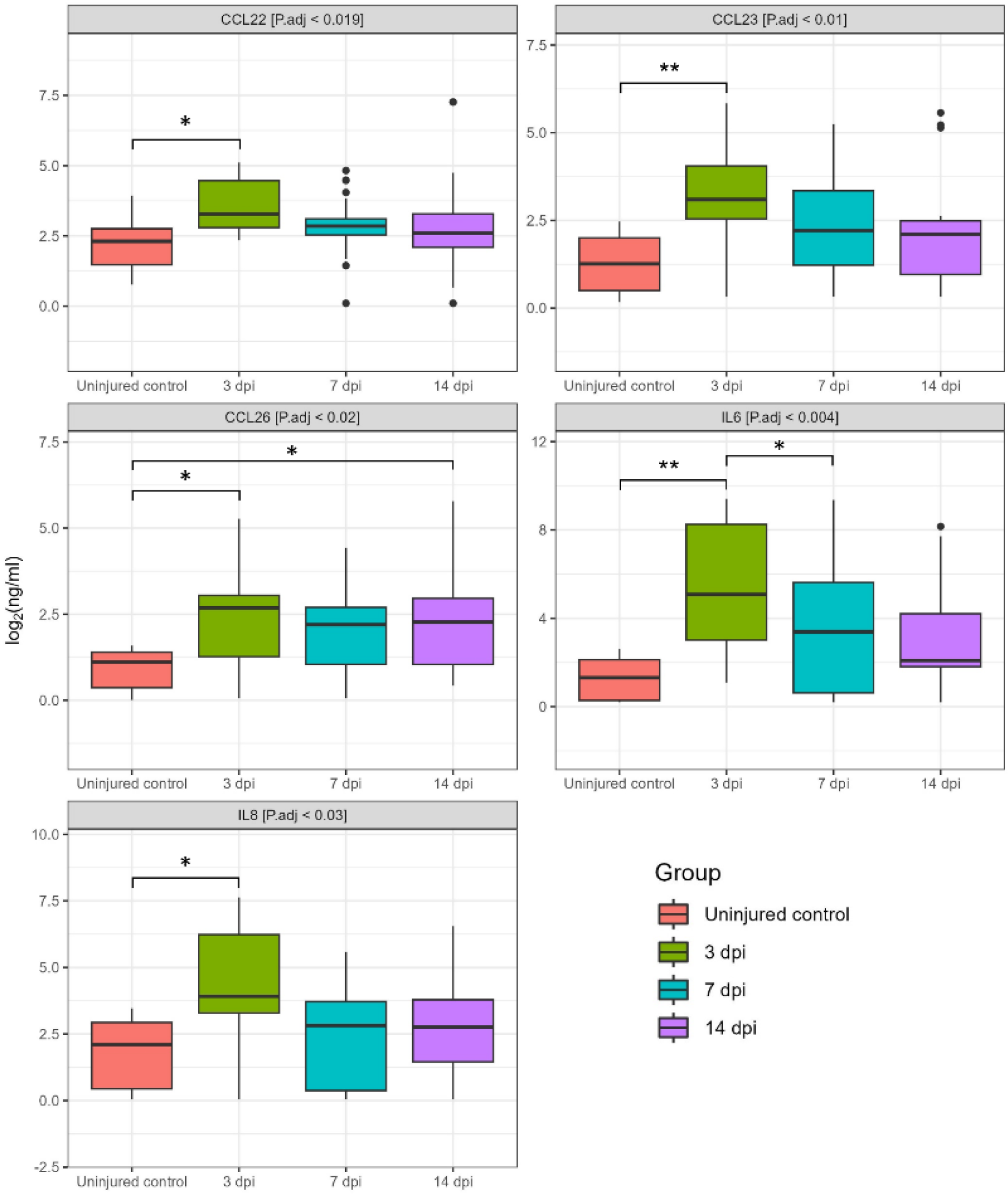
Log2-transformed CSF cytokine concentrations (ng/mL) between 3 dpi (green columns, n=15), 7 dpi (blue columns, n=21), 14 dpi (purple columns, n=23) patients and uninjured subjects (red columns, n =10). *P.adj < 0.05, **P.adj < 0.01.

At 14 dpi, expression levels of CCL22 (1.3-fold), IL-8 (2-fold), IL-6 (2.6-fold), CCL23 (2.8-fold) and CCL26 (4.2-fold) were also increased compared to uninjured control samples. However, it is important to note that abovementioned changes were only statistically significant for CCL26 [SCI 8.69 (1.83–18.32) vs. uninjured control 2.06 (0.44–3.06), P.adj <0.012]. In addition, we found a significant difference in IL-6 levels between the 3 and 7 dpi groups with a downward trend as the post-traumatic period increases.

We segregated cytokine levels based on the region of injury for determination whether the region of SCI affects CSF cytokine levels. There were no discernible differences in cytokine levels among patients with varying SCI regions and the uninjured controls at 3 dpi. However, we observed that CCL17 levels in patients with C injury were significantly elevated compared to SCI at L region and uninjured control subjects at 7 dpi, showing increases of 45-fold (P.adj <0.005) and 10-fold (P.adj <0.045), respectively (Figure 3A). SCI at Th region had CCL3 levels in 2-fold (P.adj <0.05) and 4-fold (P.adj <0.003) higher compared to SCI at C region and uninjured control subjects at 14 dpi, accordingly.

**Figure 3.**
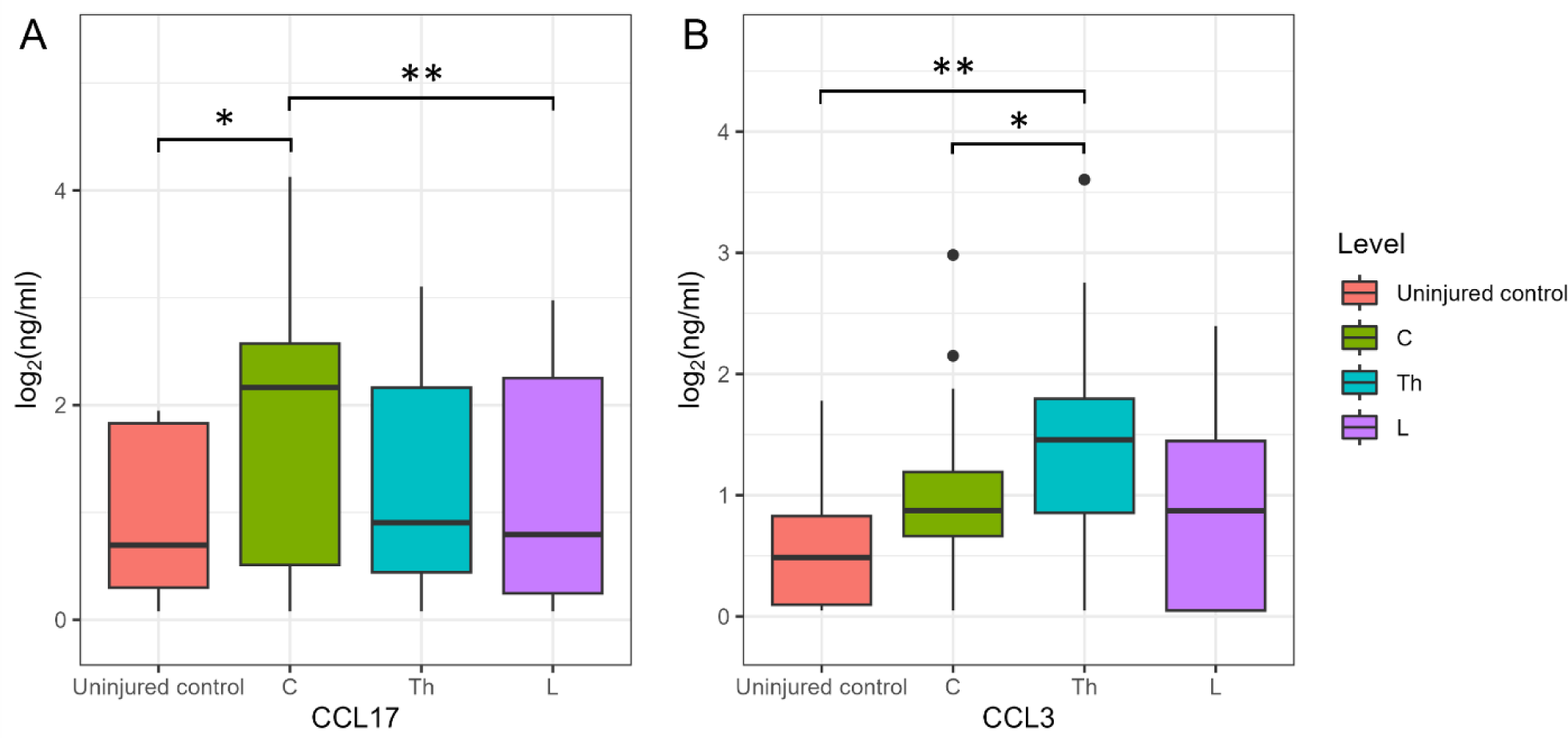
Log2-transformed CSF CCL17 and CCL3 concentrations (ng/mL) at 7 (A) and 14 (B) dpi, considering the cohort of cervical (C, n = 14), thoracic (Th, n = 11), and lumbar (L, n = 4) patients and uninjured subjects (red columns, n =10). *P.adj < 0.05, **P.adj < 0.01.

### 3.2. Dynamics of blood serum cytokine profile after spinal cord injury

Based on the analysis of blood serum, we found significant changes in cytokine profiles after SCI. Out of the 40 cytokines we analyzed at 3 and 7 dpi, the levels of CCL26, CXCL6, GMCSF, IFN-γ, IL1b, IL4, IL-8, IL10, IL16, CXCL11, CCL8, CCL7, CXCL9, CCL3, CCL19, CCL17 showed significant deviations when compared to uninjured control samples (Figure 4, Supplementary Table S1).

**Figure 4.**
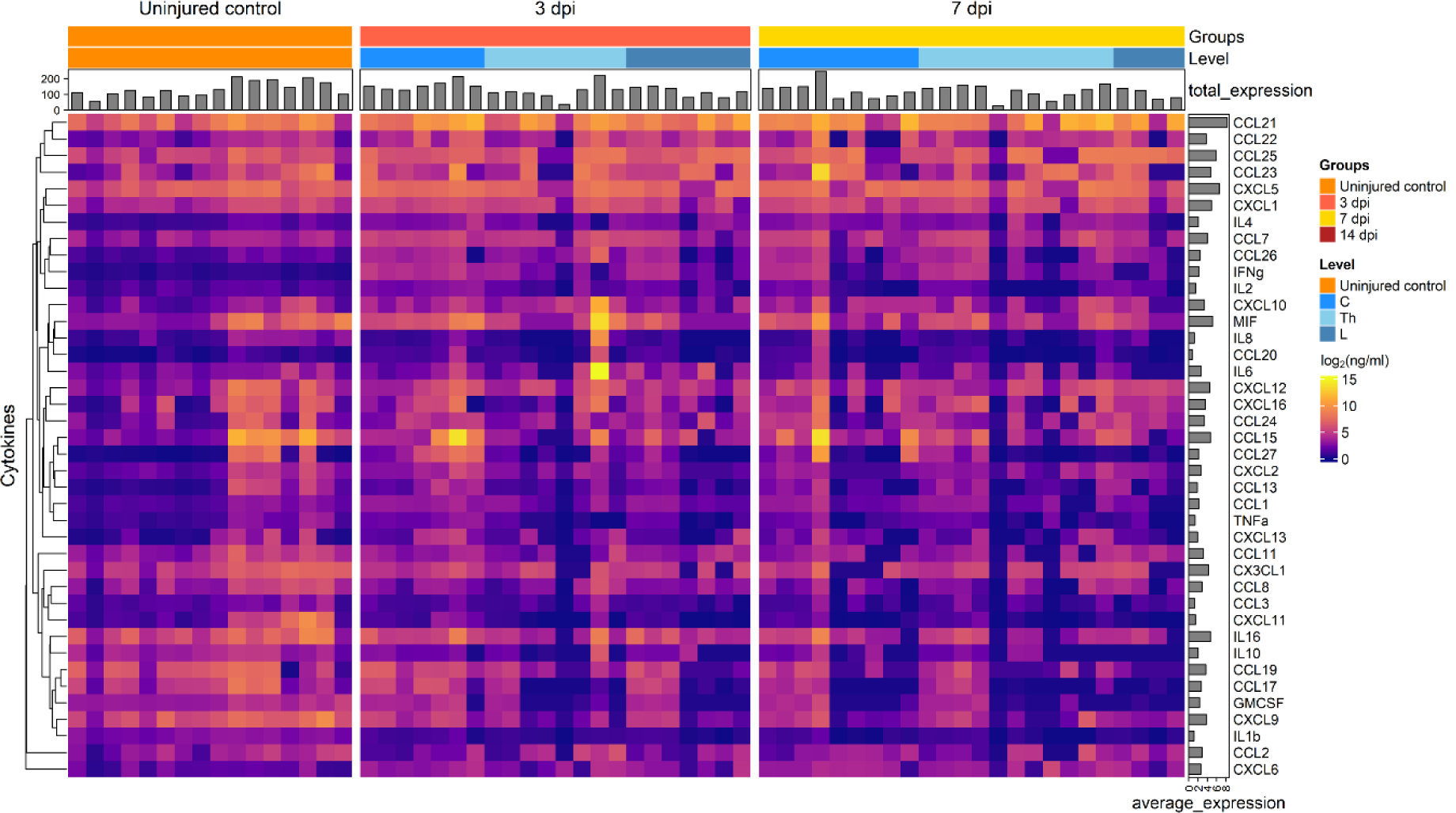
Graphical representation showing log2 cytokine concentrations (color keys), generated with the multiplex analysis of the blood serum collected at 3dpi (n = 22), 7dpi (n = 24) or from uninjured control (n = 16). A dendrogram resulting from hierarchical clustering of cytokines is shown on the left.

We found a significant increase in the levels for CXCL6 (2.8-fold, P.adj < 0.008), IL4 (4.5-fold, P.adj < 0.04), CCL7 (5.7-fold, P.adj < 0.01), CCL26 (6-fold, P.adj < 0.002) and IFN-γ (26.9-fold, P.adj < 0.0001) at 3 dpi compared to uninjured control samples (Figure 5). At 7 dpi, the levels of abovementioned cytokines also remained elevated: CXCL6 (4,2-fold, P.adj < 0.0002), IL4 (4,8-fold, P.adj < 0.0004), CCL7 (1.7-fold, P.adj < 0.014), CCL26 (5,7-fold, P.adj <0.01) and IFN-γ (14,9-fold, P.adj < 0.0005).

**Figure 5.**
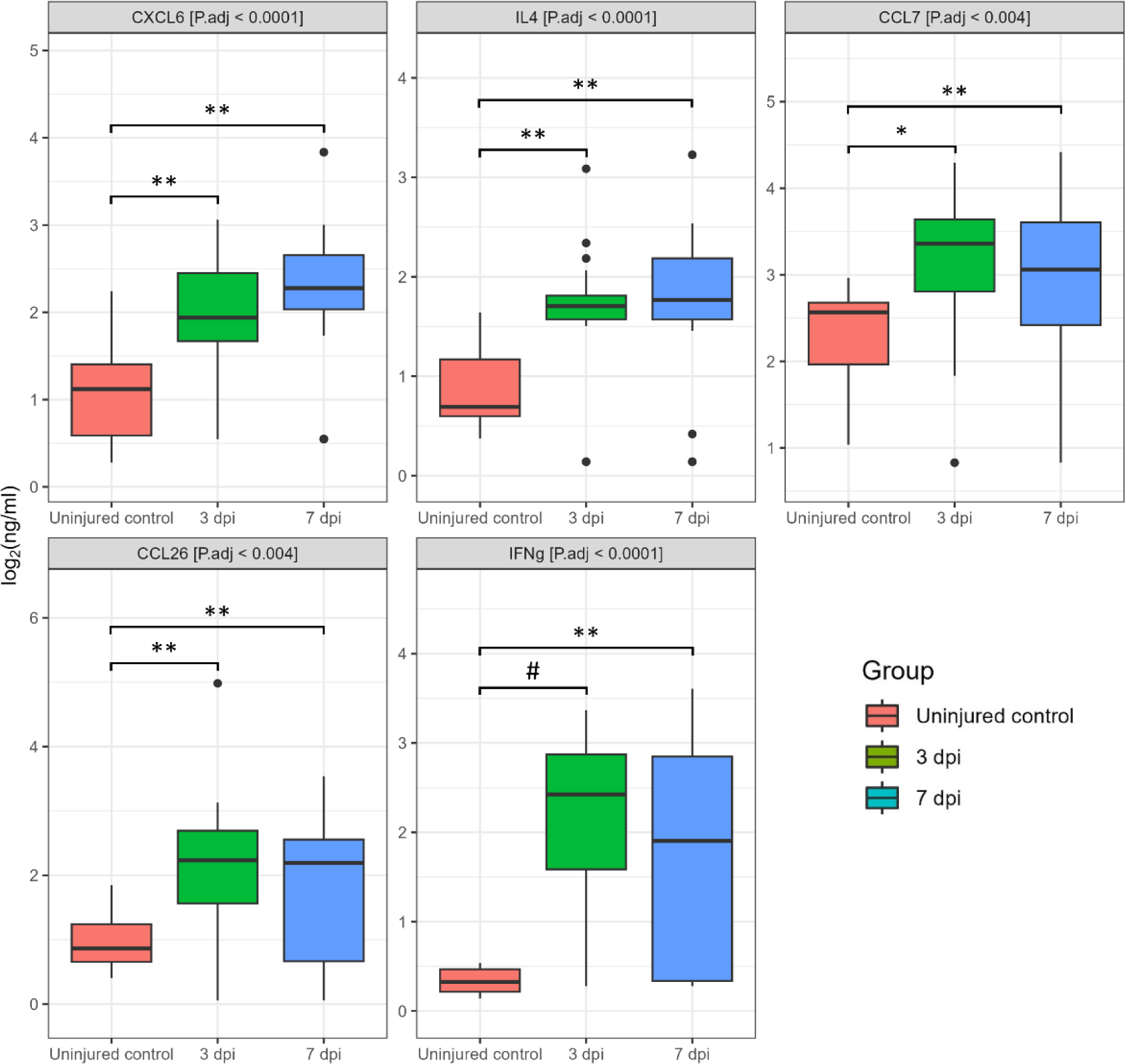
Log2-transformed blood serum cytokine concentrations (ng/mL) between 3 dpi (green columns, n=22), 7 dpi (blue columns, n=24) patients and uninjured subjects (red columns, n =16). *P.adj < 0.05, **P.adj < 0.01, #P.adj < 0.0001.

We also detected a significant decrease in several other cytokines at 3 and 7 dpi, including CCL3, CCL8, IL-8, CCL19, IL10, IL16, IL1b, CXCL9, CXCL11, GMCSF, CCL17 (Figure 6). In particular, next cytokines were lower compared to uninjured control at 3 dpi: CCL8 (2.5-fold, P.adj < 0.05), CXCL11 (2,9-fold, P.adj < 0.005), IL-8 (2,9-fold, P.adj < 0.02), IL10 (4-fold, P.adj < 0.0005), IL1b (7-fold, P.adj < 0.0001) and CXCL9 (7,4-fold, P.adj < 0.0001). At 7 dpi we continued to observe decreased level of the abovementioned cytokines: CCL8 (2.9-fold, P.adj < 0.007), CXCL11 (8,7-fold, P.adj < 0.0008), IL-8 (2,6-fold, P.adj < 0.02), IL10 (21.4-fold, P.adj < 0.0001), IL1b (10.7-fold, P.adj < 0.0001) and CXCL9 (10,7-fold, P.adj < 0.0001). In addition, CCL17 (324.5-fold, P.adj < 0.0001), GMCSF (21.2-fold, P.adj < 0.009), CCL19 (19.9-fold, P.adj < 0.003) and IL16 (4.6-fold, P.adj < 0.004) all showed a significant decrease in the SCI patient samples at 7 dpi compared to uninjured control. It is important to note that we did not observe significant differences in the blood serum cytokine profile when comparing 3 and 7 dpi.

**Figure 6.**
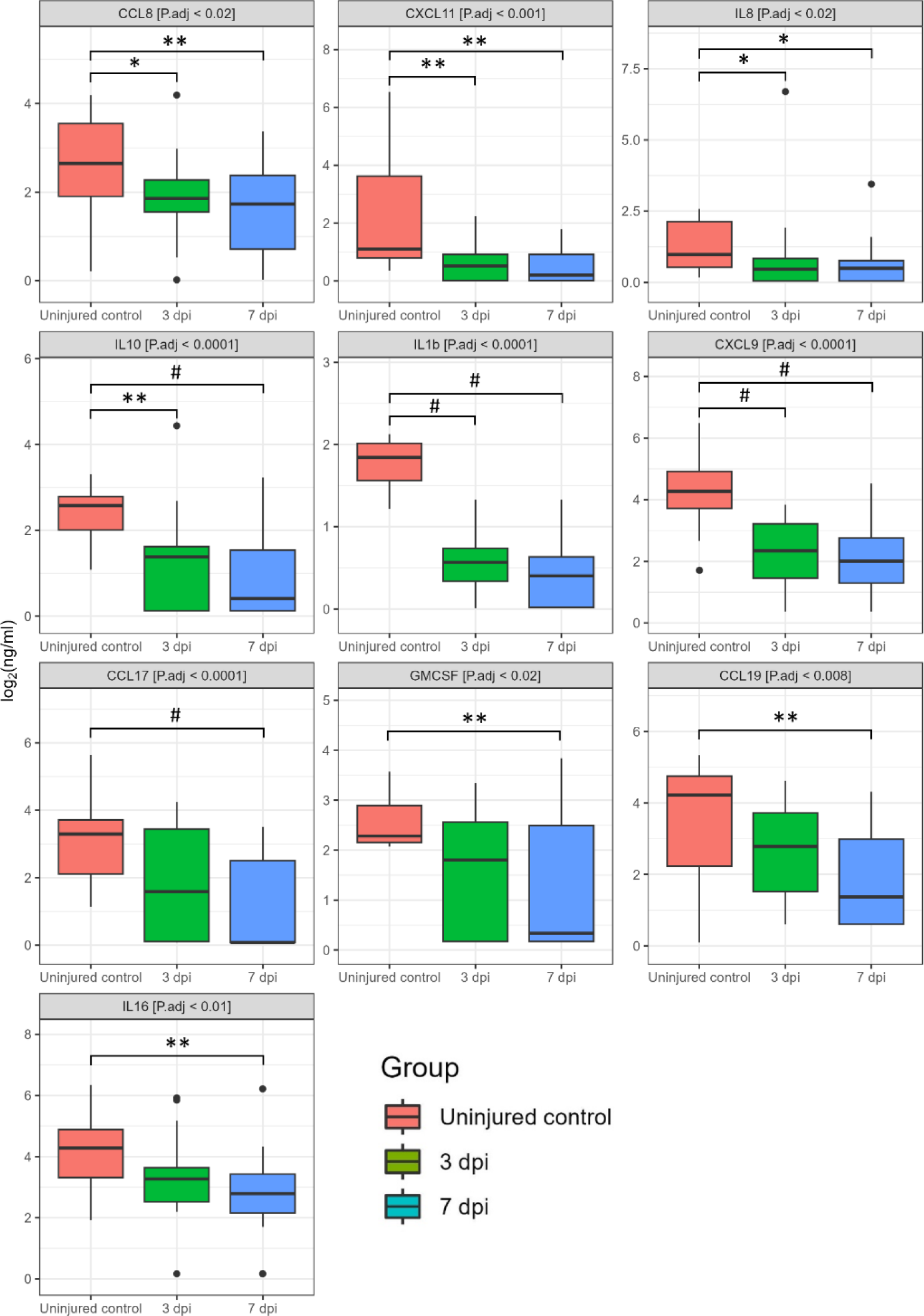
Log2-transformed blood serum cytokine concentrations (ng/mL) between 3 dpi (green columns, n=22), 7 dpi (blue columns, n=24) patients and uninjured subjects (red columns, n =16). *P.adj < 0.05, **P.adj < 0.01, #P.adj < 0.0001.

We also segregated cytokine levels based on the region of injury for determination whether the region of SCI affects blood serum cytokine levels. It is interesting to note that changes in cytokine levels associated with the region of injury were detected only at 3 dpi. The CCL27 level in patients with C injury was elevated 560-fold (P.adj <0.027) and 360-fold (P.adj <0.034) compared to SCI at L region and uninjured control subjects, accordingly (Figure 7). The concentration of IL2 was observed to significantly increase by 2.7-fold (P.adj <0.012) and 2.5-fold (P.adj <0.001) in patients with C injury compared to those with SCI at the Th region and uninjured control subjects, respectively. While the concentrations of CCL17 [0.08 (0.08–4.34)], GMCSF [0.42 (0.19–4.58)], and TNFa [0.34 (0.02–0.84)] in patients with Th injury decreased compared to SCI at C region in 566-fold, 28-fold and 10-fold (CCL17 [45.34 (13.03–55.02)], GMCSF [11.97 (10.25–16.76)], TNFa [3.41 (2.84–5.95)]) and uninjured control subjects in 76-, 20- and 3-folds (CCL17 [25.96 (7.71–39.98)], GMCSF [8.81 (7.62–17.18)], TNFa [1.12 (0.8–2.78)]).

**Figure 7.**
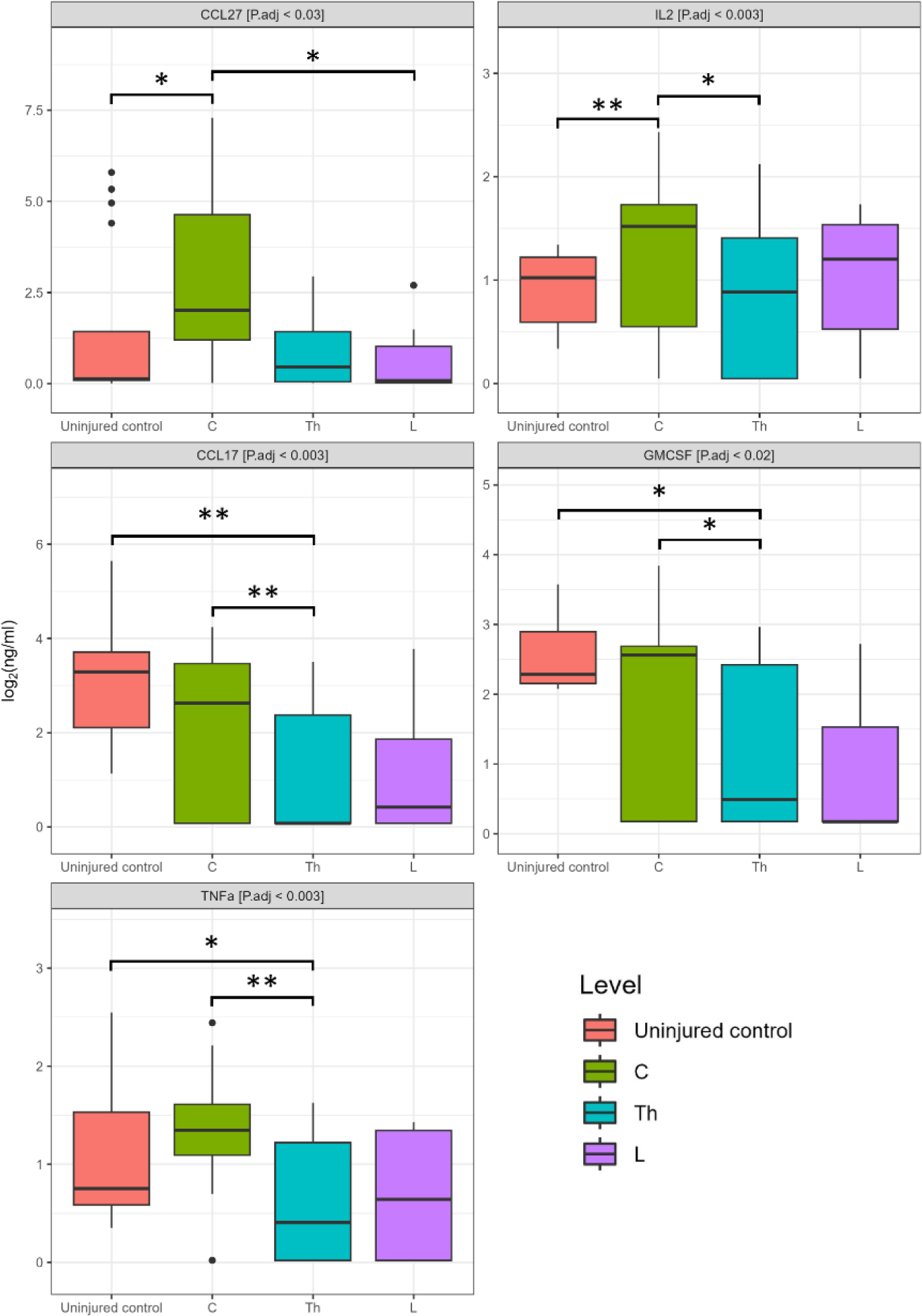
Log2-transformed blood serum cytokine concentrations (ng/mL) between patients at 3dpi injury, considering the cohort of cervical (C, n = 7), thoracic (Th, n = 8), and lumbar (L, n = 7) patients, and uninjured control subjects (n = 16). *P.adj < 0.05, **P.adj < 0.01.

## 4. Discussion

In our current study, we performed a multiplex analysis of 40 cytokines in the CSF and serum of patients with SCI at different time periods after injury (3, 7 and 14dpi), comparing the obtained data with those of uninjured control subjects. We observed both upregulation and downregulation of various cytokines after SCI.

One of the most notable results was a sustained increase in CCL26 levels in SCI in both CSF by 3 and 14 dpi and in serum by 3 and 7 dpi. These results align with our previous study, suggesting a potentially universal role for CCL26 in the inflammatory response following traumatic SCI, regardless of the severity of the injury (Ogurcov et al., 2021). Elevated levels of CCL26, both in CSF and blood serum, have been observed in numerous diseases, including Alzheimer’s Disease (Westin et al., 2012), asthma (Larose et al., 2015), rheumatoid arthritis (Zhebrun et al., 2014), cancer (X. Chen et al., 2021), and experimental autoimmune encephalomyelitis (Shou et al., 2019). This suggests that CCL26 alone might not provide a definitive diagnostic tool due to its broad involvement in inflammatory responses, it could potentially be used in conjunction with other specific biomarkers to enhance the accuracy of early diagnosis and to tailor personalized treatment strategies for SCI patients.

In our study, we observed a significant increase in IL-8 level in the CSF of patients following SCI, while a decrease in IL-8 level was noted in the blood serum of the same patient group at 3 dpi. This contrasting pattern suggests a differential regulation of IL-8 in the CNS and the systemic circulation following SCI, highlighting the complex and dynamic nature of the inflammatory and immune response to such injuries. Similar results on the level of expression of IL-8 in CSF and blood serum were observed by Kossmann et. al. (1997) examining patients with TBI. It has been suggested that IL-8 release in the CSF after neurotrauma may be associated with BBB dysfunction and nerve growth factor production (Kossmann et al., 1997).

Elevated level of IL6 was also found in the CSF of SCI patients at 3 dpi. The increase in the abovementioned cytokine is consistent with the results of a study by Kwon et al. (2010) where multiplex analysis found that IL6 levels in the CSF of SCI patients are extremely high, especially in patients with severe damage. These results highlight the important role of IL6 and IL8 in the acute inflammatory response after SCI. IL6, for example, is significantly increased within hours of injury in both rodents and humans, and blocking its receptor (IL6R) has been shown to reduce glial scar, neutrophil and macrophage invasion, and improve functional recovery after SCI (Okada et al., 2004; Nakamura et al., 2005). Using electrochemiluminescence, Fertleman et al. (2022) found that levels of many cytokines in CSF, including IL6 and IL8, are significantly increased after orthopedic surgery. The next day after the operation, their concentration even exceeds the figures recorded immediately after surgery (Fertleman et al., 2022). Based on the above data, it can be assumed that after spinal decompression surgery for SCI, the levels of these cytokines may also increase. Apparently, IL6 and IL8 alone might not provide a definitive diagnostic tool, but can be used in combined biomarker approaches.

In our study, we also observed a significant increase in CCL22 level in the CSF of patients with SCI at 3 dpi. Functionally, CCL22 is a potent chemoattractant for antigen-testing but non-resting T-lymphocytes, as well as for NK cells and monocytes. CCL22 levels in CSF have been reported to be elevated in women with multiple sclerosis (Galimberti et al., 2008). These data, combined with our observations, highlight the potential importance of CCL22 as an inflammatory marker not only in neurodegenerative diseases such as multiple sclerosis, but also in acute SCI.

We observed a sharp decrease in the pro-inflammatory cytokine IL1b at 3 and 7 dpi in blood serum, mirroring trends in our previous SCI study in rats (Mukhamedshina et al., 2017). In the study performed by Biglari et al. IL1b concentration fluctuated greatly between 4 hours and 7 dpi. However, between 1 and 4 weeks after injury, the level of IL1b in blood serum significantly decreased only in those patients in whom the improvement was less significant (Biglari et al., 2015).

A significant decrease in blood serum chemokines CXCL9 and CXCL11 was found by our investigation after SCI at 3 and 7 dpi. Three structurally related chemokines (CXCL9, CXCL10, CXCL11) form the subgroup the interferon (IFN)-inducible non-ELR CXC chemokine (Müller et al., 2010). The chemokines CXCL9 and CXCL11 function as central modulators of the immune response, predominantly in inflammatory cascades. Stein et al. (2013) in pilot study on the simultaneous assessment of the level of 21 cytokines in the blood plasma of patients with chronic (>1 year from initial injury) SCI, on the contrary, found a significant increase in CXCL9 levels in patients with chronic SCI compared with uninjured control subjects (Stein et al., 2013).

We also found that the level of IFN-γ in the blood serum of patients was significantly increased by both 3 and 7 dpi, which may indicate inflammation and secondary damage that occurs after SCI (Hellenbrand et al., 2021). IFN-γ has been previously shown to be involved in antimicrobial immunity and is elevated in various bacterial diseases (Shtrichman & Samuel, 2001). This cytokine is also known for its role in antitumor immunity (Kursunel & Esendagli, 2016). Monitoring of this cytokine can provide insight into the severity of injury and the body’s response. However, since the level of IFN-γ is also increased in other pathological conditions, its use will only be justified along with other specific biomarkers.

Our data confirm that the inflammatory and immune response to SCI is a complex and multifaceted process involving various molecules. Unfortunately, the assessment of cytokines or chemokines levels separately cannot be a valuable tool for diagnosing and monitoring the status of patients with SCI. That’s why combined biomarker approach could offer a more comprehensive understanding of the individual patient’s inflammatory profile, thereby improving the precision of therapeutic interventions. Despite the increased activity of researchers in this area, additional investigations is needed to better understand the role of injury-responsive cytokines in the context of SCI and their potential application in clinical practice.

## 5. Study Limitations

Although our study provides valuable insights, it is not without its shortcomings. This study was limited to just 40 patients with SCI, requiring larger sample sizes in future studies. The limited number of participants likely reduced the study’s ability to detect significant cytokine patterns. We encountered challenges in simultaneously collecting blood and cerebrospinal fluid from all SCI patients, attributed to cerebrospinal fluid dynamics disorders arising in certain posttraumatic phases. It should also be noted the need to confirm the results using other methods of molecular analysis.

## 6. Conclusions

In light of the need to find more reliable and stable SCI biomarkers, we conducted this study, which greatly expands on our previous analysis. In an earlier published paper, we identified several cytokines in blood serum that could potentially serve as biomarkers for the severity of SCI (Ogurcov et al., 2021). However, given the active involvement of cytokines in the regulation of the inflammatory and immune response and their possible crucial importance for the outcome of SCI (Hellenbrand et al., 2021), we hypothesized that analysis of blood serum alone may not fully reflect the actual activity of cytokines at the site of injury in the CNS. In this context, in the current study, we extended our analysis to cytokine profiles in both CSF and blood serum in more patients and at various stages after SCI. This allowed us to explore in more detail the dynamics of change and the relationship between cytokine levels, injury severity, and recovery time, paving the way for a better understanding of the pathophysiology of SCI and potentially more targeted and personalized therapeutic interventions.

## Author Contributions

Conceptualization, S.O. and Y.M.; methodology, E.G.; software, D.S.; validation, Y.M., D.S. and A.R.; formal analysis, I.K., D.S., A.S. and S.O.; investigation, S.O., I.S., D.S., A.S. and Y.M.; resources, A.R.; data curation, A.R.; writing—original draft preparation, Y.M., I.K., S.O. and D.S.; writing—review and editing, A.R.; visualization, S.O., I.S. and D.S.; supervision, A.R.; project administration, Y.M.; funding acquisition, A.R. All authors have read and agreed to the published version of the manuscript.

## Funding

The study was funded by the subsidy allocated to Kazan Federal University for the state assignment № FZSM-2023-0011 in the sphere of scientific activities.

## Acknowledgments

This paper has been supported by the Kazan Federal University Strategic Academic Leadership Program (PRIORITY-2030).

## Institutional Review Board Statement

The study was conducted according to the guidelines of the Declaration of Helsinki, and approved by the Kazan Federal University Local Ethical Committee (Protocol No. 3, 23 March 2017).

## Informed Consent Statement

Informed consent was obtained from all subjects involved in the study.

## Data Availability Statement

The data presented in this study are available on request from the corresponding author. The data are not publicly available due to the evolving nature of the project.

## Conflicts of Interest

The authors declare that there are no conflict of interest regarding the publication of this paper.

## Notes

### Competing Interest Statement

The authors have declared no competing interest.

### Funding Statement

The study was funded by the subsidy allocated to Kazan Federal University for the state assign-ment № FZSM-2023-0011 in the sphere of scientific activities. This paper has been supported by the Kazan Federal University Strategic Academic Leadership Program (PRIORITY-2030).

### Author Declarations

This study was approved by the Kazan Federal University Lo-cal Ethical Committee (Protocol No. 3, 23 March 2017). Written informed consent was obtained from each subject before CSF and blood serum samples were obtained.

