## Supplementary Table 1 for "Comparative Analysis of Cytokine Profiles in Cerebrospinal Fluid and Blood Serum in Patients with Acute and Subacute Spinal Cord Injury"

**Supplementary Table S1.** Cytokine concentrations (ng/mL) in CSF and blood serum at 3, 7 and 14 days post-spinal cord injury in patients and uninjured control subjects.

| Cytokine | CSF | | | | Blood serum | | |
| --- | --- | --- | --- | --- | --- | --- | --- |
|  | Uninjured Control | 3 dpi | 7 dpi | 14 dpi | Uninjured Control | 3 dpi | 7 dpi |
| CCL26 | 1.83 (1.49)  2.06 (0.44–3.06) | 26.53 (49.22)  13.58 (2.95–20.28) * | 12.71 (18.73)  7.98 (1.83–13.83) * | 29.89 (71.25)  8.69 (1.83–18.32) | 2.00 (1.46)  1.38 (0.93–2.46) | 14.87 (29.67)  8.34 (3.77–13.77) ** | 9.49 (9.47)  7.97 (0.97–11.88) * |
| IL8 | 85.76 (58.74)  7.23 (0.73–19.03) | 394.96 (608.69)  49.05 (25.82–606.05) * | 45.90 (74.71)  15.77 (0.46–40.11) | 85.05 (176.30)  14.86 (3.39–42.92) | 3.92 (3.91)  1.72 (0.69–7.43) | 37.58 (171.92)  0.59 (0.05–1.34) * | 2.06 (6.13)  0.64 (0.05–1.15) * |
| IL6 | 770.85 (308.25)  2.72 (0.34–7.47) | 3299.32 (5184.97)  159.82 (20.36–6097) ** | 802.66 (2543.30)  28.66 (0.87–275.19) | 385.09 (890.10)  7.05 (5.10–66.34) | 6.54 (7.19)  5.03 (2.45–5.61) | 4595.94 (21495.77)  5.84 (3.32–11.32) | 6.15 (9.12)  1.70 (0.22–7.33) |
| CCL23 | 3.79 (3.63)  2.54 (0.73–6.46) | 58.16 (91.97)  21.05 (11.69–56.93) ** | 33.85 (52.82)  8.12 (2.40–27.35) | 31.76 (70.31)  7.17 (1.61–11.04) | 74.37 (148.79)  15.56 (6.23–77.32) | 114.60 (190.44)  28.73 (11.06–168.27) | 474.01 (1991.08)  13.78 (7.39–122.53) |
| CCL22 | 13.67 (15.44)  9.38 (3.39–14.78) | 54.74 (49.80)  25.37 (15.43–88.21) * | 25.78 (30.02)  16.30 (11.54–21.33) | 84.57 (293.49)  12.44 (7.19–25.98) | 10.41 (9.80)  7.10 (3.43–14.99) | 36.36 (41.16)  15.31 (9.28–51.76) | 42.83 (107.43)  16.63 (8.46–23.18) |
| IL1b | 0.51 (0.44)  0.18 (0.10–0.80) | 1.47 (2.04)  0.71 (0.31–1.50) | 0.59 (0.64)  0.47 (0.11–0.79) | 1.15 (2.12)  0.68 (0.11–0.96) | 5.08 (1.67)  5.33 (3.77–6.51) | 0.78 (0.62)  0.76 (0.40–1.09) # | 0.59 (0.64)  0.50 (0.02–0.88) # |
| CXCL9 | 42.46 (64.80)  5.21 (2.18–59.83) | 22.73 (27.37)  7.24 (3.62–35.97) | 12.52 (12.89)  7.74 (2.65–19.68) | 48.43 (156.97)  5.54 (2.33–13.18) | 114.44 (154.44)  70.53 (40.30–136.71) | 14.99 (14.80)  9.45 (3.28–24.02) # | 15.48 (23.09)  6.57 (2.65–14.88) # |
| IFN-γ | 11.31 (5.11)  8.79 (3.71–11.77) | 17.21 (17.34)  14.55 (3.92–25.39) | 12.60 (9.19)  13.82 (8.26–16.27) | 17.10 (10.70)  15.13 (8.50–24.89) | 0.42 (0.20)  0.39 (0.24–0.60) | 10.78 (8.55)  10.36 (3.90–16.69) # | 9.40 (10.13)  5.76 (0.40–16.27) ** |
| IL10 | 3.61 (3.32)  1.73 (0.13–3.64) | 18.06 (31.71)  9.59 (4.21–13.60) | 6.04 (9.52)  2.74 (1.00–7.65) | 5.83 (7.82)  2.91 (0.64–7.36) | 12.40 (7.81)  12.14 (6.47–15.23) | 6.45 (17.49)  2.98 (0.13–4.07) ** | 3.15 (5.28)  0.57 (0.13–3.70) # |
| CXCL6 | 10.16 (11.95)  3.24 (0.73–18.61) | 10.61 (7.26)  11.86 (5.68–13.90) | 6.66 (9.04)  5.74 (0.73–7.47) | 31.12 (86.38)  6.19 (2.70–9.41) | 2.61 (2.25)  2.07 (0.80–3.11) | 7.62 (5.79)  5.95 (4.32–10.59) ** | 10.31 (9.01)  8.75 (6.67–13.21) ** |
| CXCL11 | 0.45 (0.49)  0.35 (0.04–0.65) | 1.31 (1.27)  0.93 (0.42–2.04) | 0.99 (1.11)  0.77 (0.01–1.02) | 2.25 (4.29)  0.74 (0.28–1.72) | 72.70 (175.50)  2.02 (1.21–36.61) | 1.56 (2.41)  0.68 (0.01–1.51) ** | 0.92 (1.33)  0.23 (0.01–1.51) ** |
| IL4 | 3.44 (2.08)  1.49 (0.58–5.03) | 6.27 (5.05)  6.20 (2.56–9.03) | 3.37 (2.92)  3.50 (0.15–4.87) | 5.14 (4.29)  4.36 (2.35–5.99) | 1.58 (1.21)  1.00 (0.82–2.26) | 5.00 (4.29)  4.50 (3.81–5.11) * | 6.08 (5.04)  4.86 (3.81–7.89) ** |
| CCL7 | 16.75 (13.56)  18.92 (2.59–28.49) | 57.68 (88.45)  33.15 (12.75–58.73) | 63.76 (86.88)  33.15 (24.61–62.72) | 37.07 (22.39)  35.78 (27.26–51.57) | 10.28 (5.16)  12.02 (6.15–13.56) | 27.90 (17.13)  27.78 (15.86–37.12) ** | 26.32 (22.44)  20.38 (10.29–35.92) * |
| CCL3 | 1.13 (1.51)  0.64 (0.11–1.30) | 4.40 (8.90)  2.01 (0.72–3.27) | 2.77 (3.68)  1.11 (0.74–3.65) | 3.52 (4.26)  1.99 (1.15–3.43 | 3.45 (3.65)  2.19 (1.51–3.95) | 1.80 (2.72)  0.99 (0.79–1.37) * | 1.66 (2.06)  1.08 (0.46–1.69) * |
| CCL17 | 2.48 (2.61)  1.04 (0.35–5.24) | 8.96 (10.43)  2.02 (0.61–15.46) | 5.25 (5.82)  1.27 (0.60–10.33) | 8.00 (13.07)  1.50 (0.50–10.95) | 55.24 (82.44)  25.96 (7.71–39.98) | 17.47 (22.41)  5.10 (0.12–30.80) | 6.22 (9.66)  0.08 (0.08–11.25) # |
| CCL8 | 1.84 (2.00)  1.13 (0.26–2.86) | 21.46 (44.34)  4.19 (2.00–9.58) | 10.40 (16.67)  2.02 (1.10–13.12) | 37.70 (101.13)  5.11 (0.97–12.44) | 22.55 (21.68)  13.56 (5.74–34.23) | 8.84 (13.36)  5.42 (3.73–8.80) | 6.76 (7.42)  4.67 (1.18–9.77) ** |
| CCL19 | 37.07 (42.37)  25.28 (10.48–46.36) | 55.46 (46.23)  46.69 (12.32–83.93) | 102.12 (150.85)  57.89 (11.93–144.13) | 72.25 (62.93)  61.54 (27.76–92.07) | 74.32 (64.38)  67.44 (8.54–114.62) | 27.17 (30.54)  15.84 (3.62–40.38) | 15.05 (21.11)  3.39 (0.84–18.98) ** |
| IL16 | 35.71 (16.60)  15.86 (8.70–82.28) | 99.85 (160.59)  31.05 (20.36–90.12) | 37.29 (35.50)  27.14 (17.44–56.47) | 47.55 (63.34)  24.76 (14.85–49.33) | 119.37 (154.81)  71.38 (27.64–131.39) | 63.89 (103.36)  25.20 (11.42–36.90) | 39.12 (99.61)  15.25 (7.65–29.64) ** |
| GMCSF | 4.47 (11.94)  0.45 (0.19–3.51) | 14.86 (17.74)  8.37 (1.18–20.88) | 9.70 (10.92)  4.18 (0.19–16.60) | 13.26 (15.60)  3.81 (1.80–21.19) | 13.03 (8.07)  8.81 (7.62–17.18) | 7.14 (7.80)  5.70 (0.19–11.97) | 6.18 (10.36)  0.42 (0.19–11.13) ** |

* Padj < 0.05, ** Padj < 0.01, and # Padj < 0.0001 comparing to uninjured control subjects.
